# Analysis of the impact of lockdown on the reproduction number of the SARS-Cov-2 in Spain

**DOI:** 10.1101/2020.04.18.20070862

**Authors:** Alexandre Hyafil, David Moriña

## Abstract

**Objective:** The late 2019 Covid-19 disease outbreak has put the health systems of many countries to the limit of their capacity. The most affected European countries are, so far, Italy and Spain. In both countries (and others), the authorities decreed a lockdown, with local specificities. The objective of this work is to evaluate the impact of the measures undertaken in Spain to deal with the pandemic.

**Method:** We estimated the number of cases and the impact of lockdown on the reproducibility number based on the hospitalization reports up to April 15th 2020.

**Results:** The estimated number of cases shows a sharp increase until the lockdown, followed by a slowing down and then a decrease after full quarantine was implemented. Differences in the basic reproduction ratio are also very significant, dropping from de 5.89 (95% CI: 5.46-7.09) before the lockdown to 0.48 (95% CI: 0.15-1.17) afterwards.

**Conclusions:** Handling a pandemic like Covid-19 is very complex and requires quick decision making. The large differences found in the speed of propagation of the disease show us that being able to implement interventions at the earliest stage is crucial to minimise the impact of a potential infectious threat. Our work also stresses the importance of reliable up to date epidemiological data in order to accurately assess the impact of Public Health policies on viral outbreak.

## Resumen

**Objetivo:** El brote de la enfermedad Covid-19 a finales de 2019 ha puesto los sistemas de salud de muchos países al límite de su capacidad. Los países europeos más afectados son, hasta ahora, Italia y España. En ambos países (y en otros), las autoridades decretaron un confinamiento, con especificidades locales. El objetivo de este trabajo es evaluar el impacto de las medidas adoptadas en España para hacer frente a la pandemia.

**Método:** Estimamos el número de casos y el impacto del confinamiento en el número básico de reproducción según los informes de hospitalización hasta el día 15 de abril 2020.

**Resultados:** El número estimado de casos muestra un fuerte aumento hasta el bloqueo, seguido de una desaceleración y luego una disminución después de la implementación del confinamiento total. Las diferencias en el número básico de reproducción también son muy significativas, cayendo de 5.89 (95% IC: 5.46-7.09) antes del bloqueo a 0.48 (95% IC: 0.15-1.17) después.

**Conclusiones:** Gestionar una pandemia como la Covid-19 es muy complejo y requiere una rápida toma de decisiones. Las grandes diferencias encontradas en la velocidad de propagación de la enfermedad nos muestran que poder implementar intervenciones en la etapa más temprana de la misma es crucial para minimizar el impacto de una potencial amenaza. Nuestro trabajo también muestra la importancia de contar con datos epidemiológicos actualizados y confiables para evaluar con precisión el impacto de las políticas de Salud Pública en el brote.

**Palabras clave:** Covid-19, estudio de evaluación, salud pública, infecciones, virus

## 1. Introduction

By late 2019 an outbreak of Covid-19 disease -caused by SARS-Cov-2 virus-started in the region of Hubei (China), more specifically in the city of Wuhan. Since then, the disease has spread all over the world, being declared pandemic by the World Health Organization (WHO) on 2020 March 11th. The rapid propagation of the virus around the world has stressed the health systems of many countries to their limit. In Europe, the most affected countries in terms of number of detected cases, hospitalizations and deaths are, to the date, Italy and Spain. This unprecedented situation for the public health systems of these countries forced decision makers to act very quickly in order to minimize the impact of the disease and to avoid collapse. In Spain, the emergency state (*estado de alarma*) was declared on 2020 March 14th and was hardened with mandatory home confinement except for vital sectors workers (including health professionals, food supply, etc.) on March 30th. Under this situation, it is urgent to assess the efficiency of social distancing and other non-pharmaceutical interventions undertaken to control the pandemics of Covid-19.^1,2^ One of the main challenges in evaluating the impact of these actions is that data are only partially available, as many of the cases are asymptomatic or with mild symptoms,^3^ and shortage of testing kits prevent testing all patients with possible Covid-19 symptoms. Therefore, the number of cases might be severely underestimated. This issue is common in epidemiology and several methods have been recently proposed to address it (see^4,5^) under specific circumstances. Another issue is that, when no massive viral testing of the population is performed, new cases are only detected once symptoms appear, at a variable delay after contamination. This complicates the assessment of the impact of interventions to reduce/slow down the spread of a virus.

Here, we aimed at evaluating the impact of the Covid-19 related non-pharmaceutical interventions undertaken in Spain over the basic reproduction ratio *R*_0_, considering three periods of time in 2020: No intervention (until March 13^th^), emergency state (March 16-30^th^ and April 13^th^-15^th^) and mandatory confinement (March 31st to April 12^th^). We inferred retrospectively the number of contamination in each Spanish region (*Comunidad Autónoma, or CCAA*) from the patterns of hospitalizations, deaths and detected cases. Analyses were performed with data up to April 15th 2020.

## 2. Methods

We modelled the dynamics of infection with a discrete-time (Susceptible-Infected-Recovered) SIR model with time-varying *R*_0_ defined for each region *r* and day *d*. We also assumed the stock of susceptible population *S*(*r, d*) is almost constant through time at *S*(*r, d*) ≈ *N*_*r*_, (where *N*_*r*_ is the size of the total population in the region; i.e. a small percentage of the population is contaminated) as is probably valid up to now. The dynamics of infected people *I*(*r, d*) in region *r* at day *d* thus varies as:

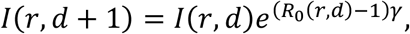

where *γ* is the recovery rate of the infection. *R*_0_ is a stochastic variable whose expected value is defined by the ongoing social distancing measures (no measure, state emergency or mandatory confinement). Those three expected values are parameters that are estimated from the reports. Formally, the priors over *R*_0_ for each region are defined as independent Gaussian Processes. The prior mean is defined by the distancing measures, while the prior covariance is Squared Exponential covariance *K* with variance σ^2^ = 0.1^2^ and length *l* = 1 day.^6^ Such prior allows to capture differences in the spread between different regions as well as temporary change of *R*_0_ within a region (Gaussian Processes enforce that these fluctuations are smooth in time). The variance term σ^2^ was set so it is unlikely that the infected population goes down in a single day by a proportion larger than the recovery rate *γ*. Based on the mean 20 days of contagious before recovery (duration of viral shedding^7^), we set *γ* = 0.05 *day*^−1^.

We assumed that the value of infected people at the beginning of the period studied (20th of February 2020) was drawn from a log-normal distribution with mean *x*_*i*_(*r*) and variance 1, where *x*_*i*_ is a parameter specific to each CCAA (estimated from the data). This variability allowed us to capture initial variations in the spread of the epidemics at the beginning of the period of study.

The number of new cases per day is *N*(*r, d*) = *I*(*r, d*) − *I*(*r, d* − 1) + *γI*(*r, d* − 1) = *I*(*r, d*) − (1 − *γ*) *I*(*r, d* − 1). This true number of cases however cannot be observed directly. Here we use a latent-state approach: we estimated the evolution of the number of true cases in each CCAA based on the recorded accumulated number of detected cases, hospitalizations and deaths provided by Instituto de Salud Carlos III^7^. We used estimates for the proportion of cases *p*_*C*_ that are detected by the Spanish Health system, the proportion of cases *p*_*H*_ that are hospitalized, and the lethality rate *p*_*D*_, as the well as the distribution of latency between infection and this three types of events *l*_*i*_(*d*) (figure 1). We used the following estimates: 15% of contaminated persons are hospitalized; 1% die from disease^8,9^ ; 30% of cases get detected (this latter number was defined arbitrarily, since the number of detected cases is roughly twice the number of hospitalizations). Note that these percentages affect the estimated number of true cases by a scaling factor, but do not affect the estimation of reproduction parameters. We used estimates of the distribution of duration of infection-to-detection, infection-to-hospitalization based on published literature^8,9^. Lauer and colleagues describe that the incubation period can be captured by log-normal distribution with median 5.1 days. To capture the extra time from the apparition of symptoms to case detection/hospitalization, we increased the median time of the log-normal distribution by 50% for case detection and 100% for hospitalization, i.e. 7.65 days and 10.2 days, respectively, while keeping the same dispersion parameter σ. For infection-to-death, we used the sum of two gamma distributions for infection-to-onset and onset-to-death with overall mean 23.9 days.^10^ Thus the expected numbers of events *y*_*i*_(*r, d*) in a certain CCAA is given by convolving the pattern of new cases with the distribution of infection-to-event.

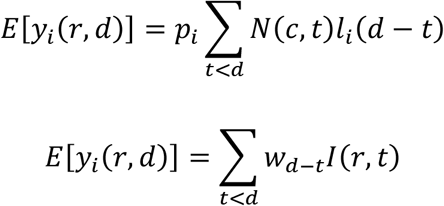

with *w*_*d*_ = *p*_*i*_(*l*_*i*_(*d*) − (1 − *γ*)*l*_*i*_(*d* − 1)). We assumed that the actual number of events recorded at that day and time was drawn from a negative binomial distribution with mean *E*[*y*_*i*_(*r, d*)] as defined above and parameter *r* = 2.^10^ The reports provide accumulated time series, so in principle new events correspond to the difference between two successive days. However new events are sometimes reported later than on the day of occurrence (especially during weekends). We estimated that 30% of events were reported on the following day, 10% two days later. Assigning this proportion of events to one or two days before they are reported allowed us to smooth the event time series. Finally, for the case of 2 CCAA (Madrid and Castilla y La Mancha, until April 12^th^ for the latter), the reported number of hospitalizations was not cumulative but corresponded to the current number of hospitalized patients related to Covid-19. For Castilla y La Mancha, we found that we could recover the reported cumulative number of hospitalizations on April 12^th^ by assuming that the duration of hospitalization is distributed uniformly between 5 and 15 days. We used this rule to estimate the cumulative number of hospitalizations for this CCAA, and applied it similarly for Madrid.

**Figure 1:**
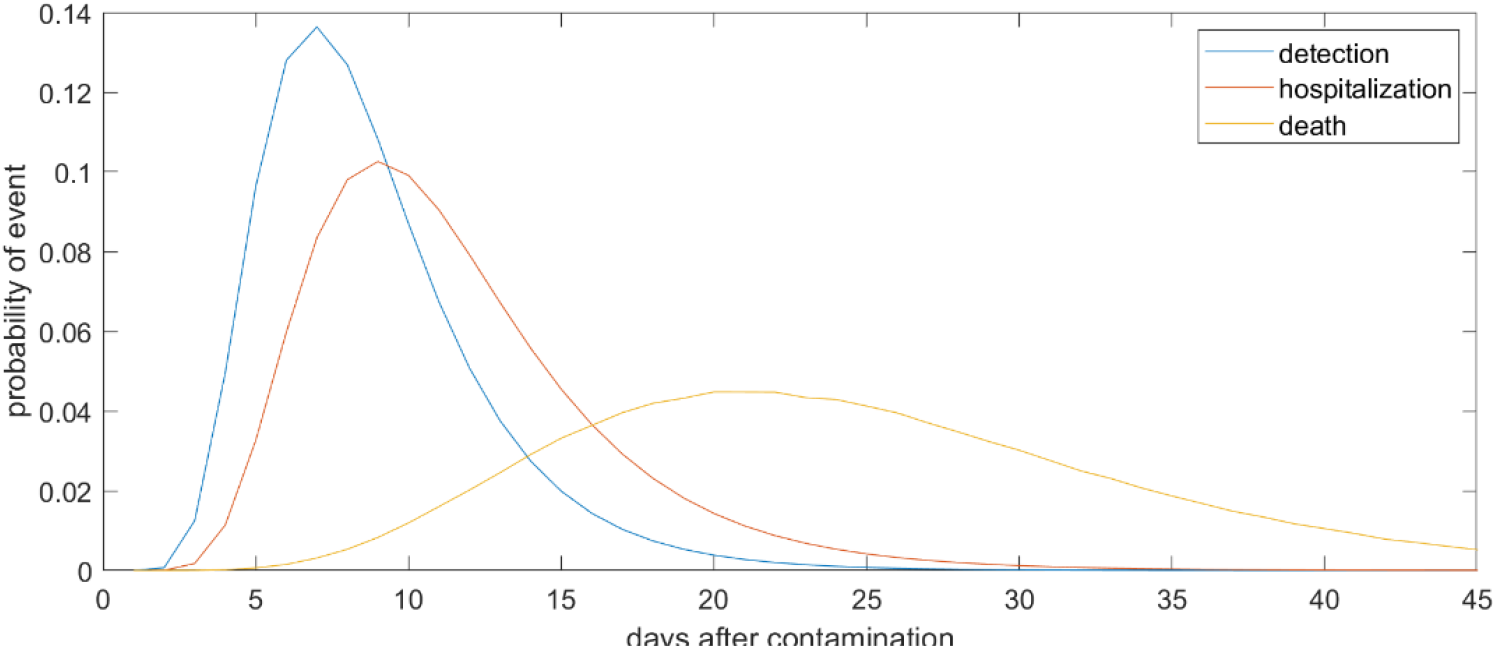
Normalized distribution for the latency from contamination to detected case, hospitalization and death. event.

There is a debate about the reliability of these data. It is believed that hospitalizations report is the most reliable of these indicators^11^, as many cases go undetected (some patients are asymptomatic or suffer mild symptoms; saturation of health systems have led to testing only more severely affected patients in some CCAA), and some deaths are not integrated in the official count because of the lack of viral charge testing. There is indeed a large variability between the fraction of hospitalizations per reported case between CCAA: it is 31% in Galicia but 82% in Comunidad de Madrid. This difference is more likely to be due to differences in detecting and reporting cases rather than in the true proportion of infected people requiring hospitalization. It is also more difficult to assess reliably the number of cases from death reports as the latency from infection to death is long and can be very variable across individuals.

Parameters of the model, including the value of *R*_0_ in the different conditions, were estimated from the data using an Expectation-Maximization algorithm (see Appendix A for details). We ran three alternative analyses: the principal analysis estimated model parameters using hospitalizations reports; two control analyses were run using either detected cases or deaths reports, which are two less reliable indicators.

## 3. Results

We present the number of Covid-19 cases in all 19 CCAA inferred from the pattern of hospitalizations in Figure 2 (see also Supplementary Figure 1 for breakdown by CCAA). The estimated cases display a sharp increase until the lockdown followed by a plateau, and then a decrease. We estimated the *R*_0_ before state of emergency, during state emergency and during enforced lockdown. *R*_0_ was found to drop from 5.89 (95% CI: 5.46-7.09) to 1.86 (95% CI: 1.10-2.63) after state of emergency, and down to 0.48 (95% CI: 0.15-1.17) after full lockdown. We estimate a total number of 0.871 million Covid-19 cases in Spain by April 15th 2020, including 0.294 active cases, 0.559 recovered and 0.018 deceased (see Figure 3 for breakdown by CCAA).

**Figure 2:**
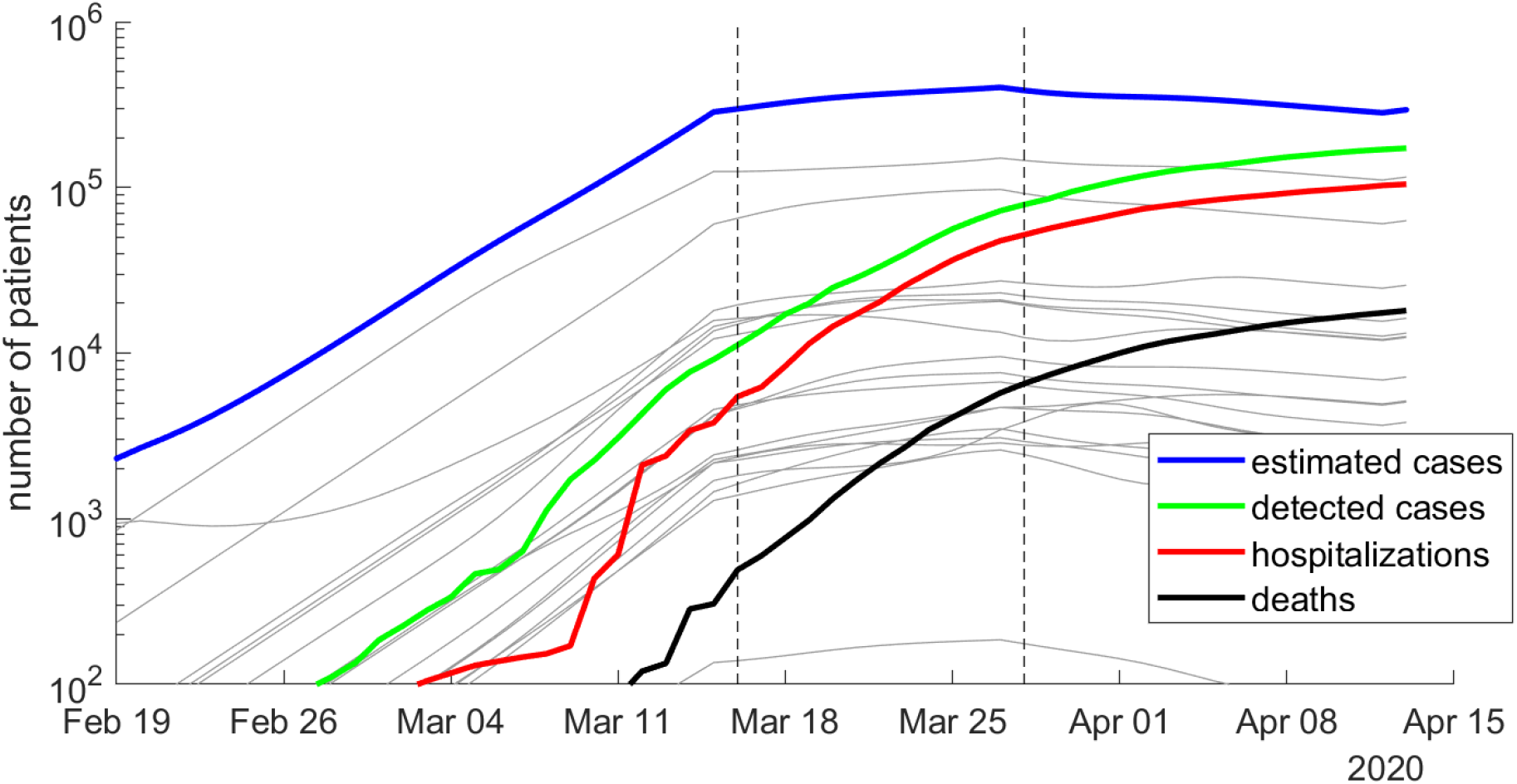
Number of estimated Covid-19 cases in Spain, inferred from the number of hospitalizations until April 15th 2020. The posterior mean of inferred cases is plotted in blue. Cumulative number of detected cases, hospitalizations and deaths are plotted in green, red and black curves, respectively. Grey curves represent the inferred number of cases in each CCAA. The onset of state of emergency and mandatory confinement are indicated by a vertical bar.

**Figure 3:**
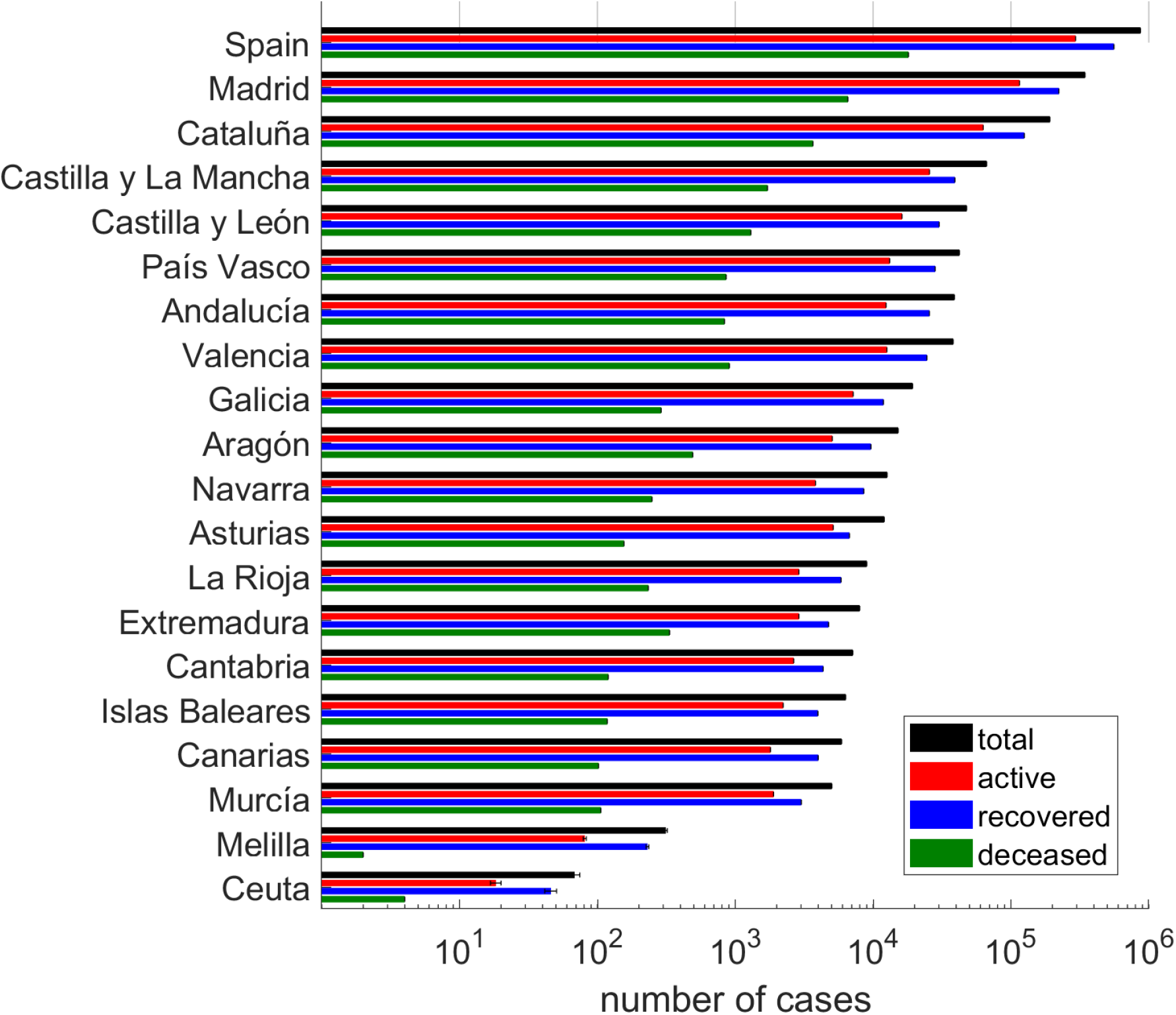
Cumulative number of cases in each CCAA, and breakdown into active cases, recovered cases and deceased patients.

Estimates were similar when we used detected cases rather than hospitalizations. Using detected case reports, we estimated the *R*_0_ to be 6.91 (95% CI: 6.75-7.39) before the state of urgency, 2.22 (95% CI: 1.92-2.74) during the state of emergency and 0.85 (95% CI: 0.5-1.05) during the full lockdown. The estimate for the cumulative number of cases was 0.823 million overall in Spain (0.351 million active).

Because deaths occur after a long and variable interval after contamination, we could not reliably estimate the value of *R*_0_ separately for the state emergency and full lockdown measures, so we simply estimated *R*_0_ before and after declaring the state of emergency. We estimated the *R*_0_ to be 6.48 before the lockdown (95% CI: 5.5-7.51), and 0.49 afterwards (95% CI: 0.16 - 1.57). The estimate for the cumulative number of cases was 2.82 millions overall in Spain (0.72 millions active).

Finally, we assessed how much our results depended on some of our assumptions, using hospitalization reports. We estimated the parameters using a different pattern of delays of notification for hospitalizations (50% reported on the same day; 20% on the next day; 20% two days later; 10% three days later). This change in delays induced a relatively small change in estimated *R*_0_ : 5.60 before the state of emergency (95% CI: 4.77-7.05), 1.66 (95% CI: 1.14-2.55) during the state of emergency and 0.60 (95% CI: 0.13-1.35) during the full lockdown. The assumed probability of hospitalizations had no impact at all on the estimated *R*_0_, and had an inversely proportional influence on the estimated number of cases: assuming 7.5% of hospitalizations instead of 15% would lead a twofold increase in the estimated number of cases.

## 4. Discussion and limitations

We found similar estimates of reproducibility number and the proportion of the Spanish population contaminated by the new coronavirus, whether they were estimated from hospitalization numbers or detected cases. Both estimates from case reports and hospitalizations suggest that only mandatory quarantine achieved *R0* lower than 1, while *R0* during state of emergency before non-essential services were shut down was estimated to be well beyond 1. This predicts that the opening of the non-essential services by April 13 may lead to a new surge of cases. Based on this empirical study, mandatory confinement is the only state-wise measure that effectively reduces the number of contaminations.

Estimates based on death reports differed considerably from those based on either hospitalizations or detected cases. This suggests that some of the assumptions and data our modeling is based on may not be accurate (although commonly used in previous studies), and shows that this can induce very large biases in the estimation of the propagation of the new coronavirus in Spain. A crucial pre-requisite for the reliability of our estimates is that the proportion of events do not vary in time. That was likely not the case neither for detected cases, as the testing policy evolved during the period of study, nor for deaths, as the saturation of health systems may have led to higher death tolls and reporting of deaths in retirement homes evolved. As the proportion of hospitalized cases is believed to be more stable across time, we believe that estimates based on the latter are more reliable than estimates based on either reported cases or deaths. Below, we further comment on the results obtained with the hospitalization reports.

Our approach is very similar to a study by an Imperial College team published last week inferring the impact of non-pharmaceutical measures (including lockdown) on propagation of the new coronavirus in 11 European countries.^10^ Both studies rely on fitting a model of infection dynamics to observed data (here hospitalizations). This contrasts with other approaches based on fitting a curve to the observed time series, (e.g. for patterns of fatalities in United States^12^ or patterns of cases in China^13^), or to model simulations studies that capture how the pattern of contacts in different scenarios (with or without social distancing measures) affect virus propagation.^14-16^ Other studies have also estimated the number of cases from the reported deaths, assuming a fixed duration from infection to death^17^.

Our modeling approach included stochasticity in the reproducibility number in each area. This notably allows to capture distinct trajectories of infection in different areas. We noted a negative correlation across CCAA between the proportion of infected people at the onset of state of emergency and the subsequent increase in infection (Figure 4, Pearson coefficient: *r* = − 0.46, *p* = 0.048). In others words, *R*_0_ at the state of emergency was smaller in the regions with more cases. This could be due to a series of factors. First, the communities with the largest proportion of cases could start developing herd immunity, hence limiting the propagation of the infection (which seems unlikely given that in the most affected CCAA, only a few percent have been infected according to our estimations). It could also be the result of local policies taken before the national lockdown in the most affected CCAA, or a better compliance of lockdown and social distancing in most affected areas. Another factor could be the migration from the most affected (especially Madrid) to less affected region before lockdown was implemented^18^, or some distortions in the reporting (under-reporting) of cases in saturated health systems.

**Figure 4:**
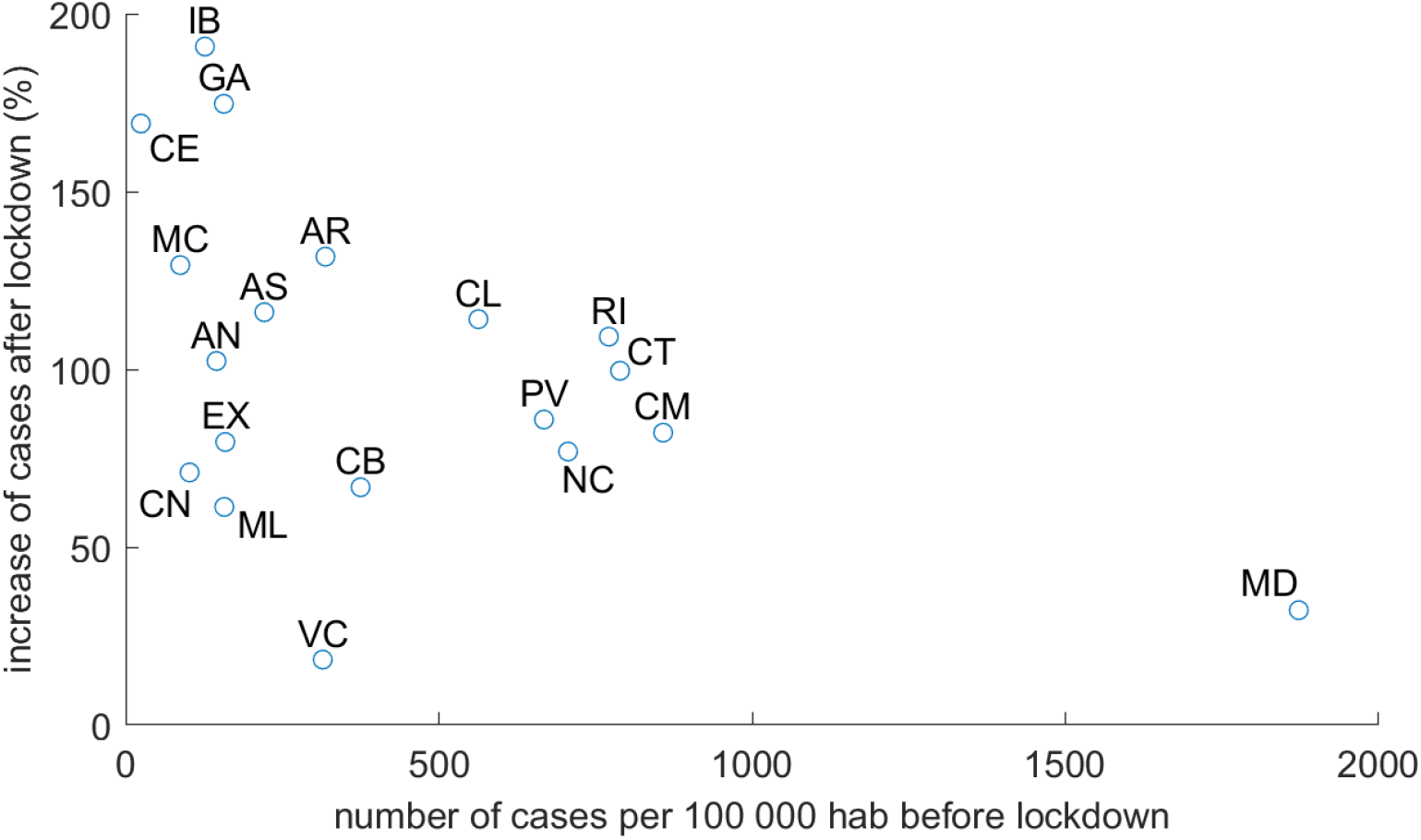
Increase in number of cases (in percent) for each CCAA in the 10 days following lockdown versus number of cases per 100.000 habitants at the time of lockdown

Our study has several important limitations. First, as noted previously, it is not clear how reliable is the data the modeling is based on: both epidemiological reports, and infectious estimate probability and latency of symptoms, case detection, hospitalization and death). It should be stressed that the unreliability of epidemiologic data (with changes of criterion of inclusion along time and between CCAA) induces important biases which impede an accurate estimation of the impact of political measures on the propagation of the new coronavirus. A faster and more reliable tracking of the epidemics could be performed if cases were reported in a systematic way dated by the onset of symptoms rather than detection, as the incubation period has been well characterized.

Second, the model captures the number of new cases each day as a proportion from the pool of infected people in the same area. It does not take into account how the age distribution in each area affects that each infection leads to hospitalizations or death, nor how the probability of infecting depends on the days from infection.^10^ Nor do it take into account mobility between regions, whose impact is believed to be more important at the initiation of the epidemics. It is also worth noting that in a locked down environment where most contacts are compartmentalized in households, it is possible that at beginning the virus continues to spread rapidly within households, but less so between households. As immunity was not taken care here, *R*_0_ may decrease significantly more without further policies, after this first wave of within-household contamination is over. Finally, we only modeled the impact of lockdown, not of other measures which were taken too close apart (banning public events, closing schools, etc.), and simultaneously in most regions, so it is not possible here to disentangle their effects precisely. The analysis also did not take into account other changes that have occurred during this period, such as measures at an individual level or at the level of companies and local institutions (usage of masks, hands washing, etc.).

In conclusion, the greatest interest should be focused on the trends in *R*_0_ found in this work, which show drastic successive reductions after the implementation of the state of urgency and forced quarantines.

## Data Availability

The study used publically available data.

## Appendix A Parameter estimation procedure

Parameters of the model *θ* include the prior mean of *R*_0_ for the three different lockdown conditions (*β*_0_, *β*_1_, *β*_2_), as well the expected value of the log-infected population at initial point *x*_*i*_(*r*). Parameters were fitted from the data by Maximum Likelihood estimation, using an Expectation-Maximization procedure.The procedure also allowed to recover the posterior distribution of true cases *p*(*I*|*y*; *θ*) for each day and CCAA. We convert the infected population to the log-scale, defining *x*(*r, d*) = *logI*(*r, d*):

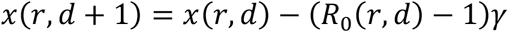

This can be turned into:

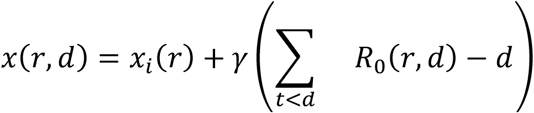

In vectorial terms, we have *x* = *T* [*x*(0, *r*), *R*_0_(*r*, :)], where *T* is an upper triangular matrix of 1 that implements the summing operation. Since both *x*_*i*_(*r*) and *R*_0_(*r*, :) have multivariate normal prior distribution, the prior over *x* is normal itself with mean *μ*_*r*_ = *T*[*x*_*i*_(*r*), *Φβ*] and covariance 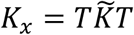 where *Ф* is an *D*-by-3 indicator matrix indicating the lockdown state for each day, and 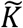 is block diagonal with submatrices 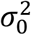 and *K*. In the Expectation step, we estimate the posterior distribution over log-infected population using a Laplace approximation *p*(*x*|*y, θ*) ≈ *N*(*m, V*). We first identified the maximum-a-posteriori variable *m* through gradient search, and then computed *V* as the inverse of the negative of the hessian joint-log-probability evaluated at *m*.

Parameters were updated in the M-step by maximizing the objective function analytically. We run the EM 10 times with different initial values for the parameters to avoid falling into local maxima of the log-likelihood. Confidence intervals for parameters were estimated using parametric bootstrapping using 20 bootstraps. All analyses were implemented in Matlab with custom codes, which will be uploaded on a public repository upon publication of the manuscript.

**Supplementary Figure 1:**
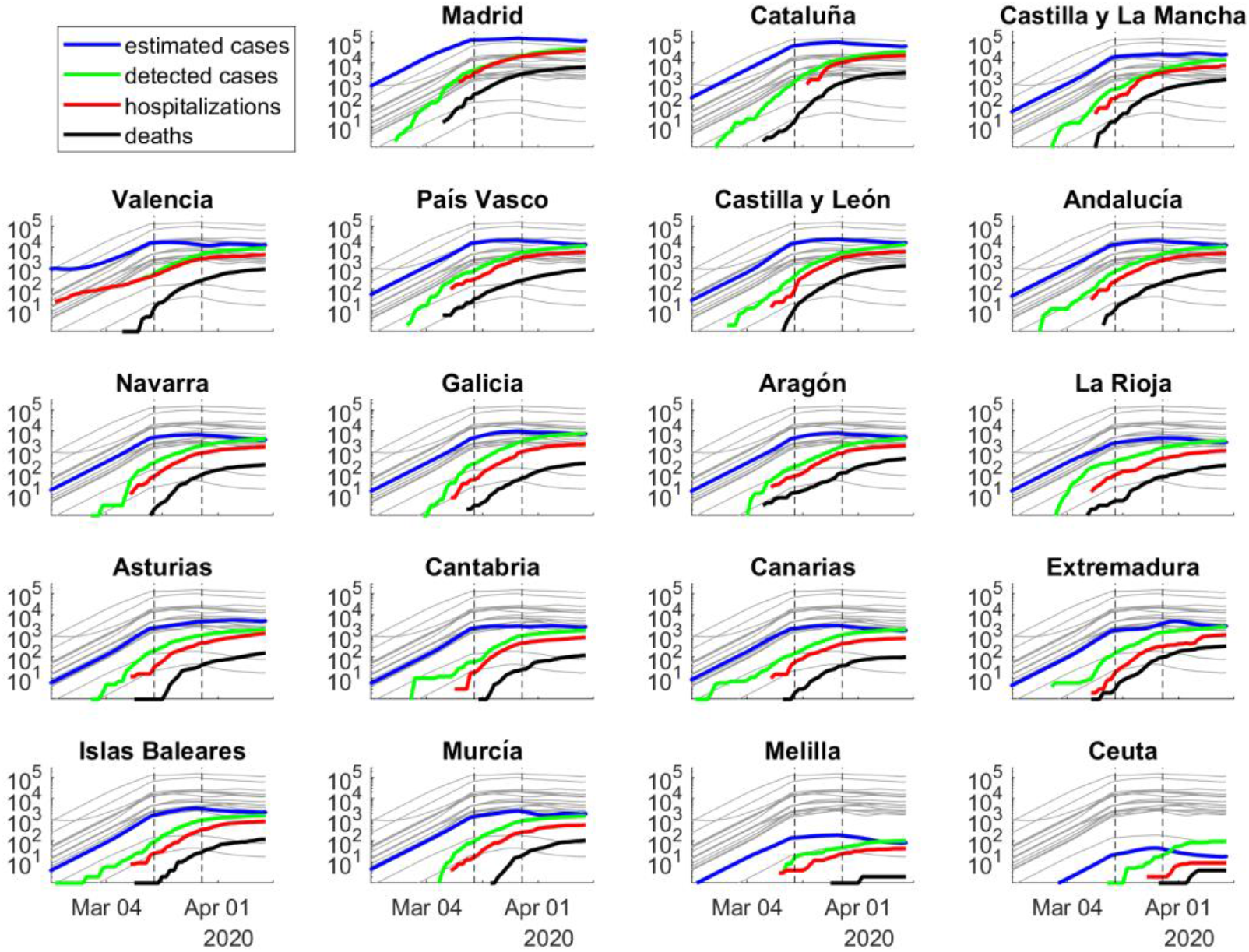
Number of estimated Covid-19 cases in all Spanish Comunidades Autónomas, inferred from the number of hospitalizations. The posterior mean of inferred cases is plotted in blue. Cumulative number of detected cases, hospitalizations and deaths are plotted in green, red and black curves, respectively. Grey curves represent the inferred number of cases in other CCAA. The onset of state of emergency and mandatory confinement are indicated by a vertical bar.

## Acknowledgements

The authors thank J. Barbosa and P. Puig for useful comments on the manuscript.

